# Continuing Medical Education Underrepresents United States’ Leading Public Health Concerns

**DOI:** 10.1101/2020.11.03.20225623

**Authors:** Nicholas A. Berry, Nicole E. Fumo, Bruce B. Berry

## Abstract

**Introduction:** Continuing medical education (CME) is beneficial to physicians in managing public health problems, yet CME courses rarely address these topics. The purpose of our study was to assess whether leading public health problems, in alignment with their burdens on society, have a proportionate amount of CME opportunities for healthcare professionals.

**Methods:** We reviewed all of the CME courses offered by the top 10 research and top 10 primary care medical schools from January 1, 2019 through June 30, 2019 for CME hours directed toward the leading public health problems: obesity, smoking, substance abuse, heart disease, COPD, lung cancer, back pain, depression, and diabetes.

**Results:** Of 9355 total CME course hours, dedicated course time, along with the number of individuals affected, and the cost to society, respectively were: obesity 118 (1.3%) hours, 93 million, $147 billion, tobacco cessation 75 (0.8%) hours, 34 million, $300 billion, and substance abuse 157 (1.7%) hours, 43 million, $300 billion.

**Discussion:** Public health problems were grossly underrepresented in the amount of dedicated course time compared to their burden on society. More CME courses offerings addressing management of the leading public health problems would likely reduce the burden of illnesses associated with those conditions.

## INTRODUCTION

Medical schools fulfill their mission, to improve the health of the community, in a variety of ways, including sponsoring continuing professional development (CME). Medicine is constantly evolving and complicated, and CME programs can help physicians stay up-to-date with changes in disease prevention and treatment. According to the Accreditation Council for Continuing Medical Education (ACCME) there were 1.2 Million (M) hours of accredited CME courses offered in 2018 at a cost of 2.8 B dollars, with medical schools receiving $367 M in CME revenue.^1^ CME topic offerings are not limited to public health problems and physicians select courses based on their own needs.

There is evidence that CME can positively impact physicians’ performance and patient outcomes in smoking cessation counseling and other conditions ^2,3^. The three most important public health concerns in the United States (US) are obesity, tobacco use, and substance abuse.^4^ The prevalence of obesity in the US is continuing to increase and now approaches 40% among adults.^5^ There are 186,000 excess deaths per year attributable to obesity at an annual cost of $147 Billion (B).^5^ Smoking tobacco has been linked to 480,000 excessive deaths every year in the US at a cost of $300 B ($8700 per smoker), with 14% of adults regularly smoking.^6^ Substance abuse, in the form of alcohol abuse, among adults is estimated to cause 88,000 deaths annually at a cost of $249 B per year and opiate abuse is linked to an additional 47,000 deaths at a cost of $78.5 B annually to society.^7,8^

The three leading causes of years of life lost (YLL) in the US are ischemic heart disease (IHD), lung cancer, and chronic obstructive pulmonary disease (COPD), and the leading causes of years lived with disability in the US are: depression, diabetes (DM), low back pain (LBP).

The purpose of our study is to determine the extent to which the top-ranked medical schools, through their CME offerings, address the leading public health problems in the US.

## METHODS

For the six-month study period starting January 1, 2019 all of the CME courses offered by US News and World Report’s top 10 ranked medical schools for research and the top 10 medical schools ranked for primary care^9^ were examined for content pertaining to the top nine public health problems: obesity, tobacco use, alcohol and drug abuse, ischemic heart disease (IHD), lung cancer, chronic obstructive pulmonary disease (COPD), low back pain (LBP), depression, and diabetes (DM)^4^. The medical school websites were reviewed monthly, and the agendas for all of their CME courses were examined. When a course was not exclusively about one of the nine key topics, only the amount of course time allocated to the study topic was credited to the topic (example: “Update on Primary Care” had 24 CME hours, but only 2 hours on obesity and 1 hour on smoking cessation, so 2 hours credited to obesity and 1 hour to smoking cessation). The same course only counted once toward the total hours, even if it was offered multiple times during the study period. Three schools were on both lists, so 17 unique schools were included in the study. To be included, a CME course had to be accredited by ACCME designee, accessible to persons outside of the institution, and an agenda had to be available so time allocated to the nine selected topics could be accurately extracted. Substance abuse was further differentiated to include opiate and alcohol use disorder. IHD did not include general topics such as congestive heart failure and atrial fibrillation or risk factor control such as hypertension and hypercholesterolemia unless they were specifically linked to an IHD agenda item.

This study was exempt from Institutional Review Board approval as all information was in the public domain. Statistical Package for the Social Sciences version 26 was used for statistical analysis.

## RESULTS

A total of 9355 CME course hours were offered. The CME instruction hours available for the three leading contributors to disability adjusted life years (DALYs) in the US were: obesity 118 (1.3%), tobacco use 75 (0.8%), and substance abuse 157 (1.7%). In the US, the adult obesity prevalence is 40%, tobacco use is prevalent in 14% of US adults, 11.2% of the US population age 12 and up reporting use of illicit drugs within the last 30 days, and alcohol use disorder (AUD) has a prevalence of 5.8% in US adults^10^, with 7 hours (0.1%) of CME course time devoted to AUD.

Course hours for the leading causes of YLL were : IHD 244 (2.6%), lung cancer 48 (0.5%), and COPD 42 (0.4%). IHD is the number one cause of death in US, and lung cancer is the leading cause of cancer deaths, COPD affects approximately 12 M people in the US.^11^

CME hours for the leading contributors to YLD were: LBP 18 (0.2%), depression 54 (0.6%), and diabetes 202 (2.2%)^4^. There are 26 M adults living with LBP, 20 M adolescents and adults experiencing a major depressive episode annually, and 34 M people living with diabetes.^12,13^ (Table 1)

**Table 1:** Burdens to United States Society for Major Public Health Problems^a^ and Corresponding Continuing Medical Education

| **CME Topic<sup>b</sup>** | **Prevalence**<br>**in US**<br>**Adults** | **CME**<br>**Hours (%**<br>**of N=9355)** | **Annual**<br>**Cost to**<br>**Society<sup>c</sup>** | **Number of**<br>**Individuals**<br>**Affected** | **Annual**<br>**Deaths** |
| --- | --- | --- | --- | --- | --- |
| Obesity | 40% | 118 (1.3%) | \$147 B | 93 M | 186,000 |
| Tobacco Use | 14% | 75 (<1%) | \$300 B | 34 M | 480,000 |
| Low Back Pain | 13% | 18 (<1%) | \$102 B | 26 M | N/A |
| Substance Abuse | 11%<sup>e</sup> | 150 (1.6%) | \$78 B | 27 M | 47,000<sup>f</sup> |
| Diabetes | 10% | 202 (2.2%) | \$327 B | 34 M | 84,000 |
| Depression | 7%<sup>g</sup>; 13%<sup>h</sup> | 54 (<1%) | \$210 B | 17 M<sup>g</sup>; 3 M<sup>h</sup> | 47,000<sup>i</sup> |
| Alcohol Use<br>Disorder | 6%<sup>g</sup>; 2%<sup>h</sup> | 7 (<1%) | \$249 B | 14 M<sup>g</sup>; 0.4 M<sup>h</sup> | 88,000 |
| COPD | 5% | 42 (<1%) | \$50 B | 12 M | 146,000 |
| Ischemic Heart<br>Disease | 3% | 244 (2.6%) | \$21 B | 8 M | 370,000 |
| Lung Cancer | <1% | 48 (<1%) | \$12 B | 0.2 M | 149,000 |

<sup>a</sup>Major public health concerns for the United States are defined in the study as each of the three leading causes for disability-adjusted life years, years of life lost, and years lived with disability.
<sup>b</sup>CME = continuing medical education; US = United States; COPD = chronic obstructive pulmonary disease.
<sup>c</sup>Cost estimates include costs of health care and lost productivity.
<sup>d</sup>Substance abuse does not include alcohol.
<sup>e</sup>Data for persons age 12 and up.
<sup>f</sup>Deaths from opioid overdose.
<sup>g</sup>Adults age 18 and older.
<sup>h</sup>Adolescents age 12 to 17.
<sup>i</sup>This is the number of suicides, which are not all from depression, but are used to estimate burden of illness.

**Table 2:** Continuing Medical Education Topic Hours Offered by Top Ranked Primary Care Medical Schools and Top Ranked Research Medical Schools^a^

| **CME Topic<sup>b</sup>** | **Primary Care**<br>**Schools CME**<br>**Hours (% of**<br>**N=2988)** | **Research**<br>**Schools CME**<br>**Hours (% of**<br>**N=7740)** | **Primary Care and**<br>**Research School**<br>**Unique Hours<sup>c</sup> (%**<br>**of N=9355)** |
| --- | --- | --- | --- |
| Primary Care | 352 (12%) | 637 (8%) | 862 (9%) |
| Neurology/Neurosurgery | 227 (8%) | 456 (6%) | 565 (6%) |
| Radiology | 188 (6%) | 452 (6%) | 471 (5%) |
| Orthopedics | 134 (4%) | 367 (5%) | 440 (5%) |
| Other<sup>d</sup> | 140 (5%) | 336 (4%) | 420 (4%) |
| Psychiatry | 120 (4%) | 308 (4%) | 418 (4%) |
| Education | 113 (4%) | 349 (5%) | 401 (4%) |
| Pediatrics | 203 (7%) | 270 (3%) | 395 (4%) |
| Cardiology | 56 (2%) | 341 (4%) | 371 (4%) |
| Gastroenterology | 95 (3%) | 326 (4%) | 360 (4%) |
| Infectious Disease | 81 (3%) | 240 (3%) | 279 (3%) |
| Oncology | 70 (2%) | 225 (3%) | 257 (3%) |
| OB/GYN | 113 (4%) | 171 (2%) | 249 (3%) |
| **Ischemic Heart Disease** | **49 (2%)** | **210 (3%)** | **244 (3%)** |
| Anesthesia | 76 (3%) | 178 (2%) | 238 (3%) |
| Urology | 79 (3%) | 189 (2%) | 238 (3%) |
| Physical Medicine | 46 (2%) | 185 (2%) | 230 (2%) |
| Pathology | 51 (2%) | 202 (3%) | 223 (2%) |
| Otolaryngology | 96 (3%) | 182 (2%) | 204 (2%) |
| **Diabetes** | **68 (2%)** | **163 (2%)** | **202 (2%)** |
| Surgery | 49 (2%) | 170 (2%) | 198 (2%) |
| Emergency Medicine | 71 (2%) | 154 (2%) | 180 (2%) |
| Hospital Medicine | 37 (1%) | 132 (2%) | 169 (2%) |
| **Substance Abuse** | **71 (2%)** | **103 (1%)** | **157 (2%)** |
| Ophthalmology | 66 (2%) | 130 (2%) | 143 (2%) |
| Leadership Development | 44 (1%) | 102 (1%) | 136 (1%) |
| Hematology | 6 (<1%) | 129 (2%) | 135 (1%) |
| Quality Improvement/Safety | 47 (2%) | 115 (1%) | 129 (1%) |
| Pulmonary/Sleep | 35 (1%) | 114 (1%) | 123 (1%) |
| **Obesity** | **27 (<1%)** | **96 (1%)** | **118 (1%)** |
| Rheumatology | 29 (<1%) | 109 (1%) | 110 (1%) |
| Endocrine | 2 (<1%) | 94 (1%) | 96 (1%) |
| Pain Management | 31 (1%) | 47 (<1%) | 79 (<1%) |
| **Tobacco Use** | **11 (<1%)** | **68 (<1%)** | **75 (<1%)** |
| LGBTQ | 44 (1%) | 45 (<1%) | 67 (<1%) |
| Peripheral Artery Disease | 17 (<1%) | 63 (<1%) | 63 (<1%) |
| **Depression** | **8 (<1%)** | **47 (<1%)** | **54 (<1%)** |
| Dermatology | 0 (0%) | 54 (<1%) | 54 (<1%) |
| Palliative Care | 16 (<1%) | 52 (<1%) | 53 (<1%) |
| **Lung Cancer** | **8 (<1%)** | **46 (<1%)** | **48 (<1%)** |
| Perioperative | 0 (0%) | 44 (<1%) | 44 (<1%) |
| **COPD/Asthma** | **12 (<1%)** | **34 (<1%)** | **42 (<1%)** |
| **Low Back Pain** | **7 (<1%)** | **12 (<1%)** | **18 (<1%)** |
| **Nine Major Public Health Topics<sup>e</sup>** | **260 (8.7%)** | **779 (10.1%)** | **958 (10.2%)** |
| **All Other Topics** | **2,728 (91.3%)** | **6,961 (89.9%)** | **8,397 (89.8%)** |

<sup>a</sup>Top 10 ranked medical schools in primary care and research per U.S. News and World Report's 2019 Best Medical School Rankings.<sup>21</sup>
<sup>b</sup>CME = continuing medical education; OB/GYN = obstetrics and gynecology; LGBTQ = lesbian, gay, bisexual, transgender, and queer; COPD = chronic obstructive pulmonary disease.
<sup>c</sup>Since three schools are counted in both the top ranked primary care and top ranked research medical schools, these three schools' CME hours were only counted once in the sum of unique hours.
<sup>d</sup>Other category contains the combined hours for topics that were offered for less than 40 hours and not pertinent to the study.
<sup>e</sup>Major public health concerns for the United States are defined in the study as each of the three leading causes for disability-adjusted life years, years of life lost, and years lived with disability. The nine topics include ischemic heart disease, diabetes, substance abuse, obesity, tobacco use, depression, lung cancer, COPD/asthma, and low back pain.

The top 10 ranked medical schools for research were more likely to offer courses addressing the leading causes of DALYs, YLL, and YLD, with 779 of 7063 total hours (10.1%) compared to the top 10 ranked medical schools for primary care offering 261 of 2988 total hours (8.7%) (p<.001 statistically significant).

## DISCUSSION

There is a large disparity between these public health concerns’ massive impacts on US health and the relatively small percentage of CME hours addressing them. Offering more CME courses on the leading contributors to DALYs, YLL, and YLD would certainly align with medical schools’ missions to improve the health of the communities they serve.

Although there were some statistically significant differences in course offerings pertaining to the 9 studied public health problems, between the top ranked research and primary care schools, the actual hours and percentages (10.1% vs 8.7%) of total hours offered were minor, and in the scope of the total offerings probably not meaningful. Making this difference even more difficult to interpret is the fact that 3 schools are included on both lists.

In this study, CME time towards tobacco cessation totaled 75 hours (0.8%) of the 9355 total hours of CME courses offered. Considering the negative impact tobacco has on the 34 M current smokers, the 50% chance of lifelong tobacco users dying due to tobacco use, and the $300 B cost to society every year^6^, it seems reasonable to have a larger share of the CME topics addressing tobacco use.

Primary care physicians readily admit they are grossly under trained in counseling techniques.^14,15^ and there are numerous studies showing the effectiveness of CME courses in educating physicians on techniques for counseling patients regarding smoking cessation ^2,3^. It seems reasonable that a renewed effort at training physicians, including the use of CME, on how to assist tobacco cessation would be very reasonable. Furthermore, reduction of tobacco use has compounded benefits on other major public health threats due to its contribution toward diseases such as substance abuse, IHD, lung cancer, and COPD.

Physicians are the customer for CME programs and as such it appears that physicians are not demanding these topics. Physicians are trained to give advice, but physicians also prefer to spend time advising patients that are more likely to actually make changes.^14,15^ Physicians may perceive many of these problems as not likely to improve and as such they are hesitant to address them.

A limitation of this study is a lack of CME course attendance numbers. This study measured the number of hours of CME offered, not the number of physicians who actually participated in each CME program. Regardless of this limitation, however, the number of CME course options is certainly less. For example, alcohol use disorder, directly affects 5.8% of US adults and 1.6% of adolescents, which is about 15 M people plus additional family and friends, but there was only one course addressing AUD out of 643 courses offered by top 10 ranked research medical schools during the study period. That one course accounted for less than one hour (0.01%) of the 7740 total CME hours offered by these medical schools.

Another limitation of our study is we reviewed only those course offerings sponsored by the top medical schools in the US. Perhaps commercial CME providers have topics more in line with public health needs. Those CME vendors were not included because their inclusion would have been arbitrary and incomplete.

## CONCLUSION

Medical schools are missing an opportunity for improving the health of the communities they serve, by offering very little CME course time addressing major public health concerns. Considering the widespread prevalence, societal cost, and disproportionally low percentage of CME hours addressing the leading contributors to premature death and years of living with a disability, change is needed in physician education. Physicians rely on medical school institutions to lead the way in healthcare advances and to provide high-quality, commercial free CME on topics that align with their missions to improve patient outcomes and community health. It is time that CME courses expand opportunities for healthcare professionals to develop additional skills, so they can reduce the burden of the most daunting public health threats to our society.

## Data Availability

All data available in the public domain and upon request to the corresponding author.

## Declarations

Institutional Review Board approval not needed, exempt research, all data in the public domain and no protected health information.

None of the authors received any payments in exchange for any work related to this study. No funding was received from any source for this study.

None of the authors have any conflicts of interest to report.

ICMJE Disclaimer completed-no conflicts

Equator research checklist consulted for “Observational Studies, topic Medical Education” no issues found.

